# Blood parameters measured on admission as predictors of outcome for COVID-19; a prospective UK cohort study

**DOI:** 10.1101/2020.06.25.20137935

**Authors:** DT Arnold, M Attwood, S Barratt, K Elvers, A Morley, A Noel, A MacGowan, NA Maskell, F Hamilton

**Affiliations:** Academic Respiratory Unit, North Bristol NHS Trust; Population Health Sciences, University of Bristol; Bristol Centre for Antimicrobial Research (BCARE), North Bristol NHS Trust; Translational Health Sciences, University of Bristol; Infection Sciences, North Bristol NHS Trust

**Author notes:** Corresponding author: David Arnold, Academic Respiratory Unit, Learning and Research Centre, Southmead Hospital, North Bristol NHS Trust, BS10 5NB.

## Abstract

**Introduction:** COVID-19 has an unpredictable clinical course so prognostic biomarkers would be invaluable when triaging patients on admission to hospital. Many biomarkers have been suggested using large observational datasets but sample timing is crucial to ensure prognostic relevance. The DISCOVER study prospectively recruited patients with COVID-19 admitted to a UK hospital and analysed a panel of putative prognostic biomarkers on the admission blood sample to identify markers of poor outcome.

**Methods:** Consecutive patients admitted to hospital with proven or clinicoradiological suspected COVID-19 were recruited. Admission bloods were extracted from the clinical laboratory. A panel of biomarkers (IL-6, suPAR, KL-6, Troponin, Ferritin, LDH, BNP, Procalcitonin) were performed in addition to routinely performed markers (CRP, neutrophils, lymphocytes, neutrophil:lymphocyte ratio). Age, NEWS score and CURB-65 were included as comparators. All biomarkers were tested in logistic regression against a composite outcome of non-invasive ventilation, intensive care admission, or death, with Area Under the Curve (AUC) figures calculated.

**Results:** 155 patients had 28-day outcomes at the time of analysis. CRP (AUC 0.51, CI:0.40-0.62), lymphocyte count (AUC 0.62, CI:0.51-0.72), and other routine markers did not predict the primary outcome. IL-6 (AUC: 0.78,0.65-0.89) and suPAR (AUC 0.77, CI: 0.66-0.85) showed some promise, but simple clinical features alone such as NEWS score (AUC: 0.74, 0.64-0.83) or age (AUC: 0.70, 0.61-0.78) performed nearly as well.

**Discussion:** Admission blood biomarkers have only moderate predictive value for predicting COVID-19 outcomes, while simple clinical features such as age and NEWS score outperform many biomarkers. IL-6 and suPAR had the best performance, and further studies should validate these biomarkers in a prospective fashion.

## Introduction

COVID-19 causes a wide spectrum of disease, from asymptomatic to severe respiratory failure. The majority of patients who present to hospital will recover but some develop rapidly progressive respiratory failure requiring ventilatory support. Biomarkers that might predict this deterioration would be invaluable when triaging patients on hospital admission to inform who can be safely discharged versus those who need careful respiratory monitoring.

The rapidly emerging literature base around biomarkers in COVID has mainly extracted data from electronic health records of large cohort studies (1-4). Far fewer have prospectively recruited and followed up patients. Those that have tested novel biomarkers have often done so at later timepoints in patients limiting extrapolation to hospital admission (5-7). Few UK studies have assessed the additional value of biomarkers compared to the routinely recorded demographic information and clinical scales (such as National Early Warning Scores or CURB-65).

The Dlagnostic and Severity markers of COVID-19 to Enable Rapid triage (DISCOVER) study prospectively recruited patients presenting with COVID-19 to a single UK hospital, with the aim of identifying the additional value of biomarkers to routine clinical care in predicting mortality, need for non-invasive ventilation or intensive care unit admission.

## Methods

This study is reported in line with the TRIPOD guidelines for prediction models (Appendix 2)

### Study design

This study aimed to assess prognostic clinical and blood biomarkers for COVID-19 disease based on the earliest available clinical and biochemical information. The primary outcome of prediction was a composite outcome of intensive care admission, non-invasive ventilation outside the intensive care unit, or death (defined below as “severe disease”).

The secondary outcome was a composite outcome of intensive care admission and death.

### Patient recruitment

All patients were recruited via the DISCOVER study, a single centre observational study at North Bristol NHS Trust recruiting patients with COVID-19, from 30.03.2020 until present (Ethics approval via South Yorks REC: 20/YH/0121, CRN approval no: 45469). Patients were recruited on the basis of a positive PCR result for SARS-CoV-2, using the established PHE assay in use at the time or a clinicoradiological diagnosis of COVID-19 disease. During the pandemic, community testing became widely available, although the results were not available to hospital staff. As such, later patients were often recruited on the basis of a history of positive testing in the community. The only exclusion criteria was an inability to consent. For patients in intensive care, family members were able to consent on behalf of them if too unwell to consent.

### Clinical information

Clinical information was recorded on a REDCap (Vanderbilt University) database (8), by the consenting nurse or physician. Routine demographics were recorded, and presence of important comorbidities. Comorbidities were defined either by their recording in the admission notes / hospital record (for hypertension, heart disease, and chronic lung disease), the presence of a positive serological result (for HIV) or for requirement for dialysis or an estimated GFR of <30ml/min (for chronic kidney disease). Ethnicity was also recorded.

The earliest admission National Early Warning Score (NEWS) was extracted from the clinical record. This is a numeric score (from 1-20), reflecting the degree of physiological dysfunction (9). Higher numbers indicate more severe physiological dysfunction. Routine biochemistry and haematology results were extracted from the clinical record, using the earliest available figure. Outcome data recorded was in line with RECOVERY, the national RCT of therapeutic interventions for COVID-19 (https://www.recoverytrial.net/). This included 28-day mortality, requirement for intensive care, ventilation, renal replacement therapy, and inotropes.

### Biomarkers and samples

For conventional biomarkers of infection (namely R-reactive protein (CRP), components of the full blood count and routine renal function) data were prospectively recorded. The first result from that admission was taken, or for established inpatients, the result on the day of the diagnosis. For other potential predictive markers (Lactate dehydrogenase (LDH), procalcitonin (PCT) interleukin-6 (IL-6), Krebs von den Lungen 6 (KL-6), Ferritin, Troponin, B-type natriuretic peptide (NT-pro BNP), soluble urokinase plasminogen activator receptor (suPAR)), analysis was performed on frozen samples in batch analysis as described below

The earliest initial sample was extracted from the blood sciences laboratory after routine testing had been performed; in admitted cases this was the initial sample taken in the Emergency Department (ED), in hospitalised cases, this was the sample from the day of diagnosis.

suPAR analysis was performed on the suparnostic ELISA platform, KL-6 and Il-6 were performed on the Fujerebio Lumipulse.

CRP, Ferritin, LDH, Troponin, NT-pro-BNP and Procalcitonin were performed on the Roche Diagnostics cobas platform. The specific analytic platforms were spectrophotometric: c701 (CRP) and c501 (LDH), immunoassay: e501 (Ferritin, Troponin, NT-pro-BNP, Procalcitonin). The full blood count was performed on the Sysmex XN (Sysmex Diagnostics).

### Statistical approach

In this study, we aimed to identify whether any individual biomarker (Lymphocyte count, Neutrophil count, Neutrophil:Lymphocyte ratio, CRP, IL-6,KL-6, suPAR, NT-pro BNP, LDH, PCT, Troponin T, Ferritin) had prognostic significance for the primary outcome as an individual marker, when used on the initial blood sample taken.

There was a deliberate focus on relatively simplistic (logistic regression) models: given the sample size, complex models would be at risk of overfitting, and the main target was to identify the additional value of a biomarker, all of which are reported on a linear scale(10). Missing data was relatively rare, and complete case analysis was performed for each individual biomarker.

For each biomarker, we performed logistic regression for each outcome. ROC curves were generated for each biomarker, and AUC figures were calculated, alongside sensitivity and specificity for each biomarker. Confidence intervals were generated around the AUC by bootstrapping.

All analysis was performed in R (version 4.0.0), using the packages “tidyverse”, “broom”, “tidymodels”, and “pROC”. Analytic code is available at: https://github.com/gushamilton/discover_prediction/

## Results

At time of writing, 182 patients have been recruited and 155 have reached 28 days post diagnosis (prespecified timing of the primary outcome). Table 1 shows the demographics of the cohort. 80 patients (55%) were male, and the median age was 60 (IQR: 46-72). 77% of the cohort had positive PCR results for SARS-CoV-2, with the remaining clinically suspected with negative testing. 85% of patients were inpatients at the time of recruitment.

**Table 1:** Demographics of the study cohort.

| Characteristic | Non-severe, N = 120<sup>1</sup> | Severe, N = 35<sup>1</sup> | p-value<sup>2</sup> |
| --- | --- | --- | --- |
| Age (18+) | 55 (44, 70) | 66 (60, 76) | <0.001 |
| Sex |  |  | >0.9 |
| Male | 65 (54%) | 20 (57%) |  |
| Female | 55 (46%) | 15 (43%) |  |
| PCR proven disease |  |  | 0.8 |
| Proven | 91 (76%) | 28 (80%) |  |
| Suspected | 29 (24%) | 7 (20%) |  |
| Status at time of consent |  |  | 0.046 |
| Inpatient | 98 (82%) | 34 (97%) |  |
| Outpatient | 22 (18%) | 1 (2.9%) |  |
| Diabetes status |  |  | 0.2 |
| No | 93 (78%) | 31 (89%) |  |
| T1DM | 2 (1.7%) | 1 (2.9%) |  |
| T2DM | 24 (20%) | 3 (8.6%) |  |
| Heart disease | 30 (25%) | 10 (29%) | 0.8 |
| Chronic Lung disease | 21 (18%) | 18 (51%) | <0.001 |
| Severe Liver disease | 2 (1.7%) | 1 (2.9%) | 0.5 |
| Severe kidney impairment<br>(eGFR< 30 or dialysis) | 11 (9.2%) | 5 (14%) | 0.4 |
| Hypertension | 29 (24%) | 12 (34%) | 0.3 |
| HIV status | 1 (0.8%) | 1 (2.9%) | 0.4 |
| Non-white | 20 (17%) | 5 (15%) | >0.9 |

<sup>1</sup> Statistics presented: median (IQR); n (%)
<sup>2</sup> Statistical tests performed: Wilcoxon rank-sum test; chi-square test of independence; Fisher's exact test

Comorbidities were relatively common within the cohort with hypertension and diabetes being the most predominant. Patients with severe disease (death, intensive care admission or non-invasive ventilation) were generally older, and more comorbid than non-severe patients. 35 patients had the primary outcome. 14 patients went to the intensive care unit, of which 4 died and 10 survived. 6 patients required NIV outside the intensive care unit, 3 of whom died.

There was a significant variation in physiological state (e.g. NEWS), with a median NEWS score of 4 (IQR 2-6), although the highest NEWS score recorded was 13. Patients that had severe disease had higher NEWS scores. There was also wide variation in functional status, with many patients having some degree of frailty, with frailer patients more likely to die or require enhanced care. Escalation status was recorded for most patients, with patients who died more likely to have limitations on ceiling of care, recorded in Table 2.

**Table 2:**
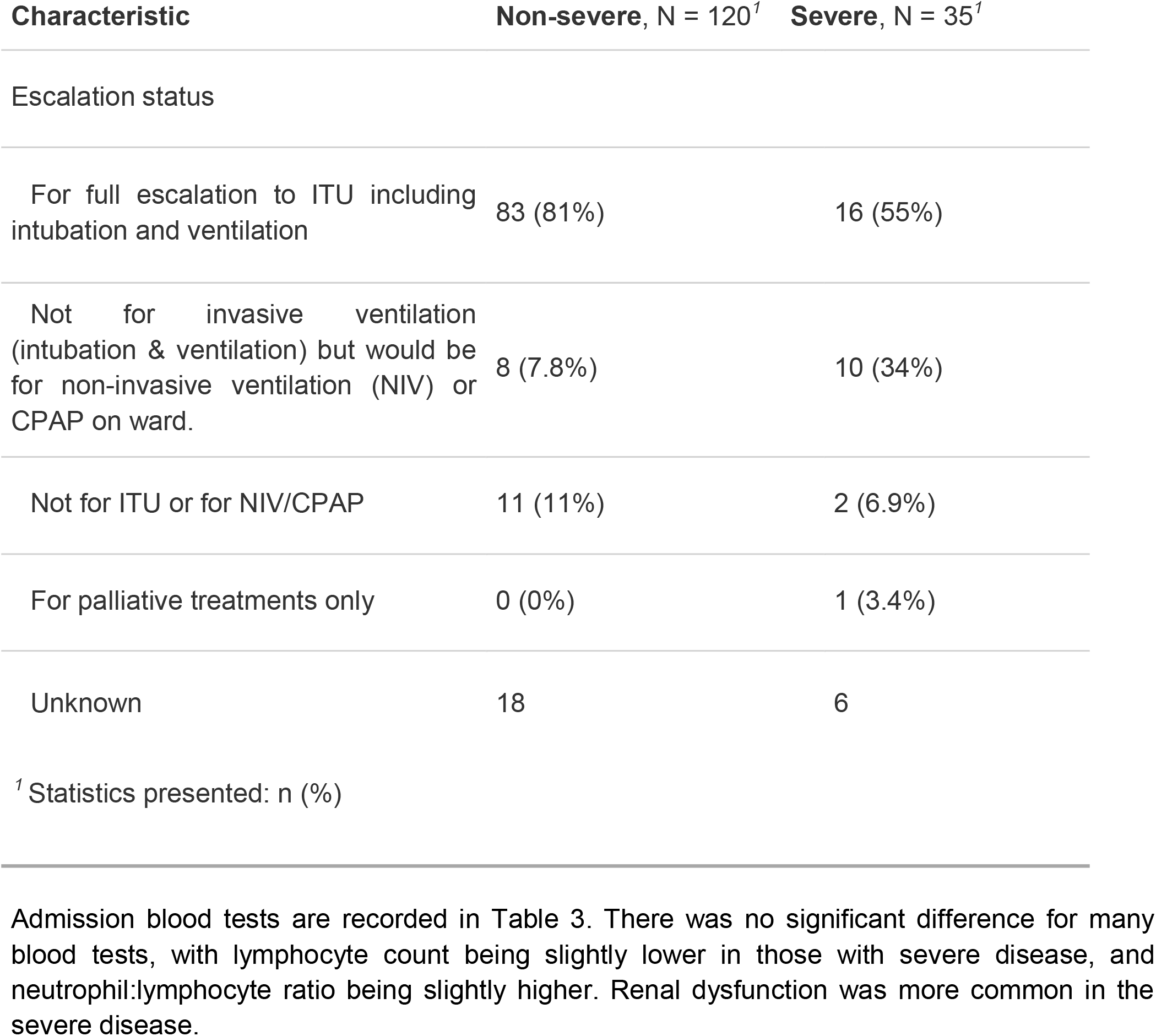
Escalation status

For each individual biomarker, box plots are shown in Figure 1 and 2. For visualisation, Ferritin, BNP, Troponin, and PCT have been logged prior to plotting to aid visualisation as they had very wide

**Figure 1:**
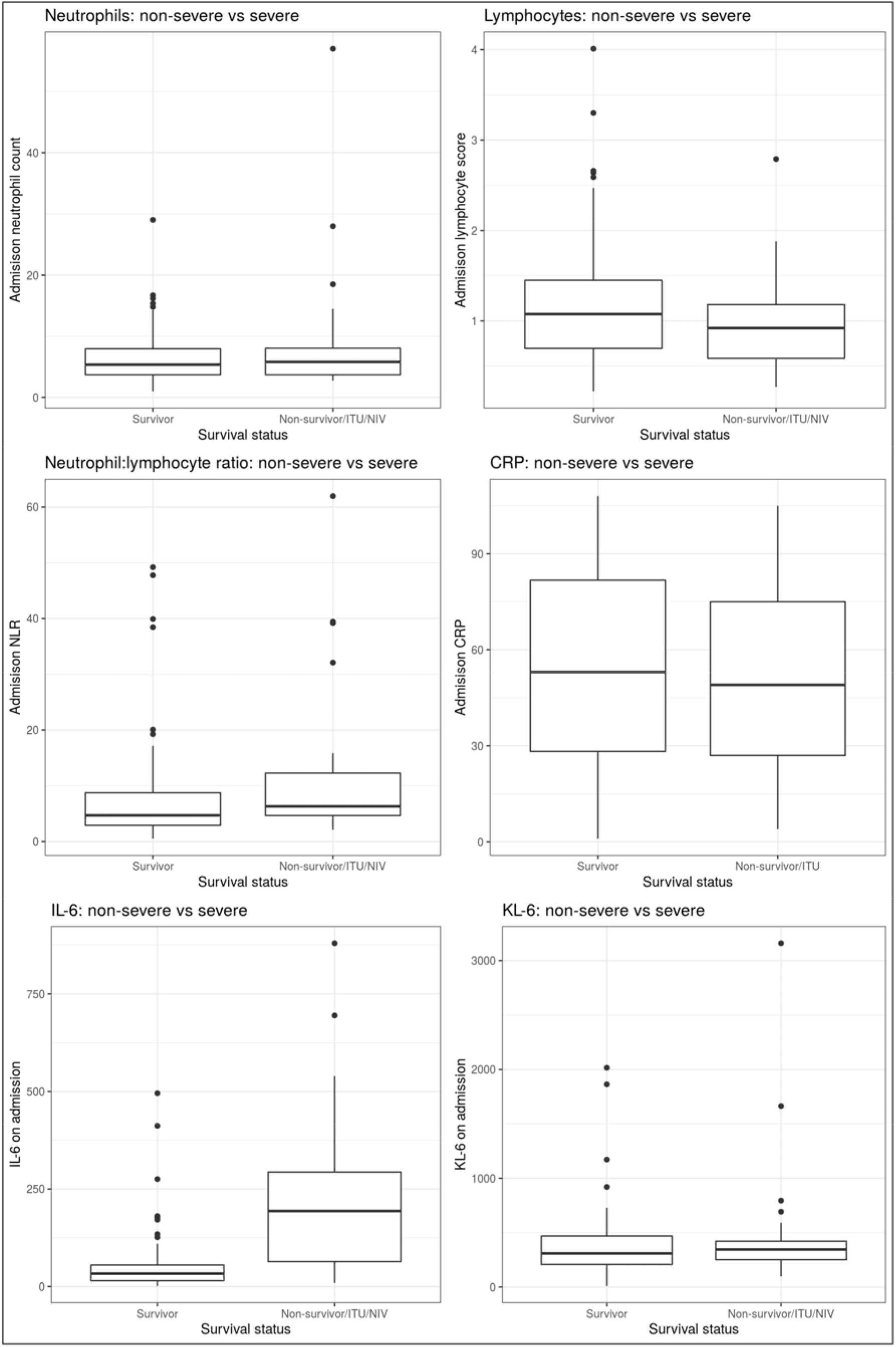
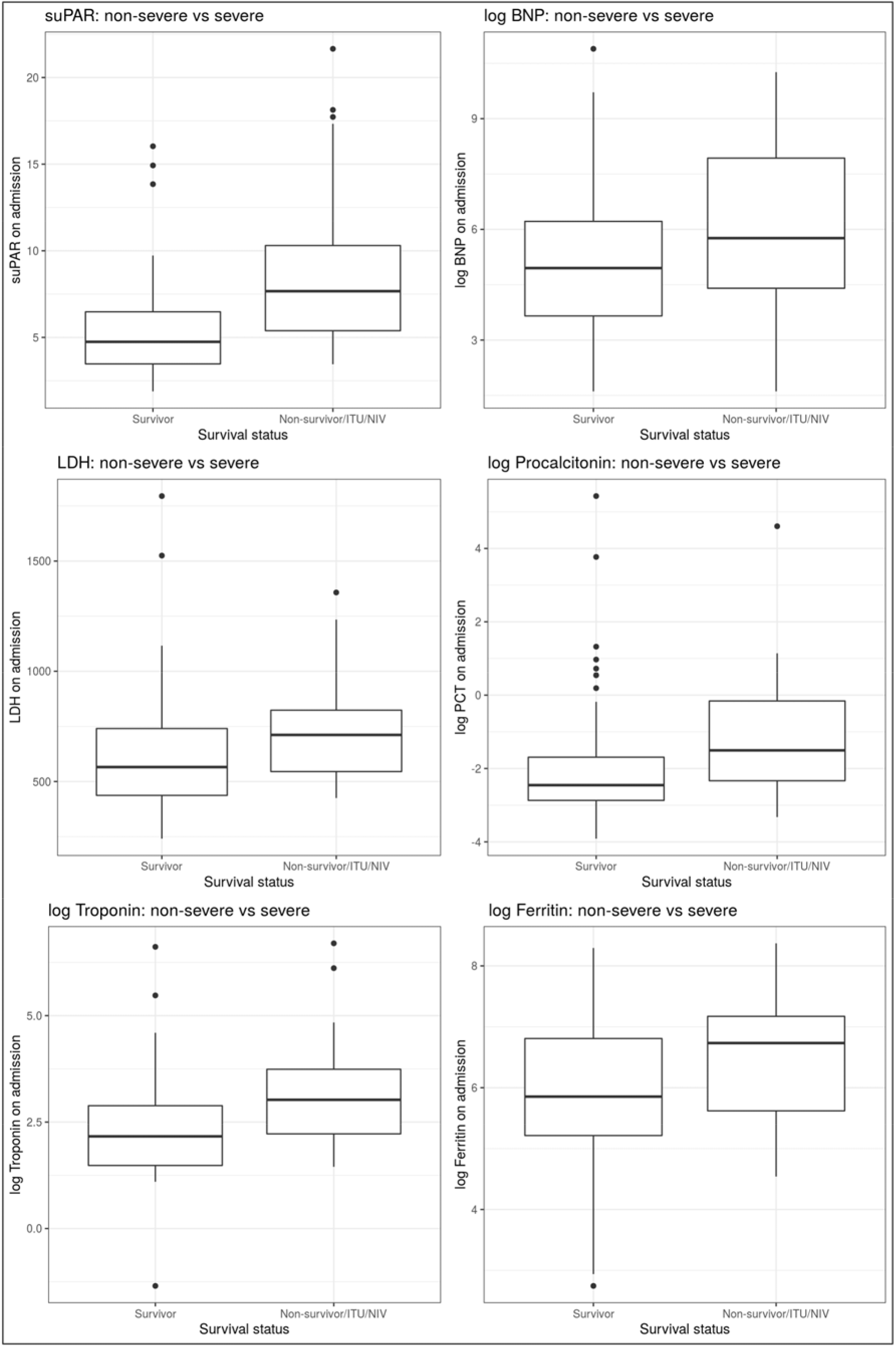
Boxplots for all biomarkers. distributions.

### Logistic regression and AUC calculation

Table 4 shows biomarker performance. Blood was not available for all participants for all tests, with the number included in each model listed. Most biomarkers had modest predictive value, with suPAR and IL-6 having the best performance (AUC 0.77 for both). Many biomarkers had negligible performance (CRP, neutrophils, lymphocytes, KL-6), with AUC figures between 0.5 and 0.6. ROC plots are available in the appendix (Figure S1) for all biomarkers. Of note, both age and NEWS score performed as well as most biomarkers.

**Table 3:**
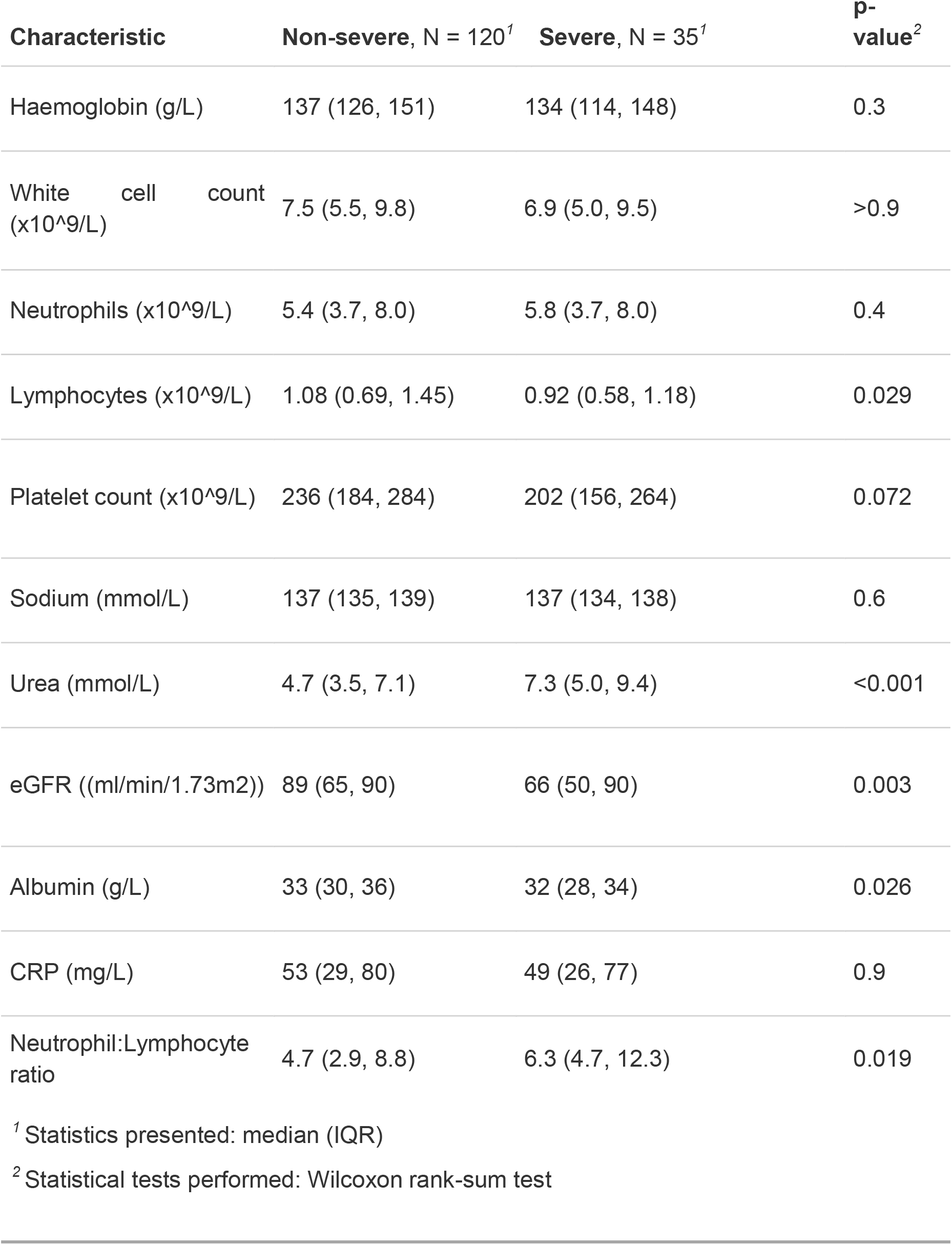
Conventional blood tests

| Characteristic | Non-severe, N = 120<sup>1</sup> | Severe, N = 35<sup>1</sup> | p-value<sup>2</sup> |
| --- | --- | --- | --- |
| Haemoglobin (g/L) | 137 (126, 151) | 134 (114, 148) | 0.3 |
| White cell count (x10<sup>9</sup>/L) | 7.5 (5.5, 9.8) | 6.9 (5.0, 9.5) | >0.9 |
| Neutrophils (x10<sup>9</sup>/L) | 5.4 (3.7, 8.0) | 5.8 (3.7, 8.0) | 0.4 |
| Lymphocytes (x10<sup>9</sup>/L) | 1.08 (0.69, 1.45) | 0.92 (0.58, 1.18) | 0.029 |
| Platelet count (x10<sup>9</sup>/L) | 236 (184, 284) | 202 (156, 264) | 0.072 |
| Sodium (mmol/L) | 137 (135, 139) | 137 (134, 138) | 0.6 |
| Urea (mmol/L) | 4.7 (3.5, 7.1) | 7.3 (5.0, 9.4) | <0.001 |
| eGFR ((ml/min/1.73m<sup>2</sup>)) | 89 (65, 90) | 66 (50, 90) | 0.003 |
| Albumin (g/L) | 33 (30, 36) | 32 (28, 34) | 0.026 |
| CRP (mg/L) | 53 (29, 80) | 49 (26, 77) | 0.9 |
| Neutrophil:Lymphocyte ratio | 4.7 (2.9, 8.8) | 6.3 (4.7, 12.3) | 0.019 |

<sup>1</sup> Statistics presented: median (IQR)<sup>2</sup> Statistical tests performed: Wilcoxon rank-sum test

**Table 4:** Biomarker performance:

| **Biomarker** | **AUC (confidence intervals)** | **Sensitivity at Youden's index** | **Specificity at Youden's index** |
| --- | --- | --- | --- |
| CRP (n = 155) | 0.52 (0.4-0.62) | 0.63 | 0.57 |
| Neutrophils (n = 153) | 0.54 (0.43-0.65) | 0.77 | 0.42 |
| Lymphocytes (n = 153) | 0.62 (0.51-0.72) | 0.69 | 0.61 |
| NLR (n = 153) | 0.62 (0.52-0.73) | 0.8 | 0.5 |
| IL-6 (n = 111) | 0.77 (0.63-0.89) | 0.75 | 0.85 |
| KL-6 (n = 108) | 0.56 (0.43-0.69) | 0.86 | 0.45 |
| suPAR | 0.77 (0.67-0.86) | 0.87 | 0.63 |
| Procalcitonin (n = 125) | 0.71 (0.6-0.81) | 0.76 | 0.67 |
| Ferritin (n = 126) | 0.65 (0.54-0.76) | 0.67 | 0.66 |
| BNP (n = 126) | 0.65 (0.53-0.76) | 0.62 | 0.74 |
| LDH (n = 126) | 0.70 (0.59-0.79) | 0.87 | 0.63 |
| Troponin (n = 126) | 0.70 (0.59-0.8) | 0.63 | 0.57 |
| NEWS score (n = 153) | 0.75 (0.65-0.85) | 0.67 | 0.66 |
| CURB-65 (n = 151) | 0.63 (0.54 – 0.73) | 0.57 | 0.73 |
| Age ( n = 155) | 0.70 (0.61-0.78) | 0.62 | 0.74 |

### Secondary outcomes

For the composite outcome of ITU and mortality, results are reported in Table S1 in the Appendix (Table S2). Due to a small number of events (15 deaths, 10 ITU admissions), confidence intervals were wider, but results were similar. Again, suPAR and IL-6 were the best performing biomarkers (AUC 0.72 for suPAR, AUC 0.70 for IL-6), with CRP having no prognostic value (AUC 0.53)

## Discussion

COVID-19 remains a clinical challenge. The majority of patients who present to hospital will recover but some develop rapidly progressive respiratory failure. Biomarkers that might predict this deterioration would be invaluable when triaging patients on hospital admission to inform who can be safely discharged versus those who might need intensive care support in the near future. This paper presents the first, prospectively recruited, UK cohort of patients with COVID-19 with targeted biomarker sampling at presentation.

### Previous literature

A large observational study recruiting from the majority of NHS hospitals (ISARIC) estimates that, of the 34608 patients with outcome data, 7374 (17%) required admission to intensive care and 11659 (33.7%) died within follow up (11). There have been numerous studies on the reasons for respiratory decline in COVID-19 pneumonia with the development of a ‘cytokine storm’ in specific patients cited as a major determinant. As a result there has been a focus on biomarkers that rise in other similar conditions such as ferritin, lactate dehydrogenase (LDH), IL-6, and soluble urokinase plasminogen activator receptor (suPAR)(12-15). A rise in cardiovascular events and coagulopathies has also been seen in patients with COVID-19 pneumonia so biomarkers such as troponin, NT-pro BNP, fibrinogen and D-dimer have been studied(16-19). Given that mortality from COVID-19 increases with age and frailty, biomarkers of frailty have been shown to be related to worse outcomes (albumin, eGFR) (2, 20). Finally, given this is a respiratory infection, blood-based biomarkers that prognosticate bacterial pneumonia have been included in treatment guidelines and even entry criteria for clinical trials, including lymphopenia, neutrophilia, procalcitonin and C-reactive protein (19, 21, 22).

### Study findings

In this study conventionally performed blood biomarkers did not predict outcome when performed on admission. Neutrophilia and C-reactive protein had AUC close to 0.5, with lymphopenia and NLR having only marginally better discriminative value. Cardiac markers were on average slightly higher in patients with worse clinical outcomes but should not be relied on to make treatment decisions at baseline. Literature suggests that they may have more utility when measured serially, especially in the very unwell patient (16).

Markers of immune activation had more promise at the admission timepoint with IL-6 and suPAR the best-performing within this cohort. It has been hypothesized that an exaggerated immune response or ‘cytokine storm’ plays a significant role in COVID-19 and several therapies immunomodulatory therapies are being trialed. IL-6 is a pro-inflammatory mediator for the induction of the acute phase response and shown to be a predictive marker of deterioration in other serious lung pathologies (23). Specific to COVID-19, Han and colleagues assessed the cytokine profile of 102 patients, admitted to a single hospital in Wuhan, by disease severity on admission. From a panel of 6 cytokines (TNF-α, IFN-γ, IL-2, IL-4, IL-6 and IL-10), serum IL-6 was predictive of current disease severity with an AUC of 0.84, although longer term outcomes were not reported(24). Another prospective study of 89 patient admitted to a German hospital demonstrated that admission IL-6 was superior to other blood-based biomarkers at predicting the need for mechanical ventilation (using a cut-off of 35pg/ml). In this study, the admission IL-6 (at a cut-off of 76.4pg/ml) had a sensitivity of 0.75 and specificity of 0.85 (AUC 0.77).

suPAR, a marker of immune activation and has been shown to predict deterioration in several infectious and inflammatory disorders. It forms part of the fibrinolysis cascade and increased levels have been shown to pre-dispose to clotting abnormality and renal dysfunction, both of which are important drivers of morbidity and mortality in COVID-19 (25). A prospective study of 57 patients presenting with COVID-19 demonstrated that admission levels of suPAR were significantly greater among patients who eventually required ventilatory support (12). A cut-off of 6ng/ml had a sensitivity of 86% and specificity of 92%. In this study it performed reasonably compared to other blood biomarkers but in our study a cut-off of 6ng/dl only had a sensitivity of 63% and specificity of 68%, with lower cutoffs increasing sensitivity at relatively little cost.

### Clinical markers

The focus of this study was the additional role of blood biomarkers when initially assessing patients presenting with COVID-19. The DISCOVER cohort also collected routinely recorded clinical data including demographics, baseline observations and initial radiography. It is notable that this easily accessible information outperformed many of the blood-based biomarkers tested.

The National Early Warning Score (NEWS) is used throughout the United Kingdom. There is currently limited evidence supporting its use in COVID-19(26-28). In China, Liao and colleagues created a non-validated COVID specific NEWS score (where age over 65 added 3 points) (29). We could not find any UK-based studies accessing the utility of NEWS in COVID-19 admission. In the DISCOVER cohort, NEWS score was as predictive as suPAR and IL6 with an AUC of 0.75 (0.65-0.85).

### Strengths

The DISCOVER cohort was prospectively recruited and attempted to analyse the earliest available clinical sample, which is a key strength. Clinical meta-data was robustly recorded, and novel biomarkers were performed in batch assays, maximising replicability and reducing bias as results could not influence patient care. Unlike other datasets (e.g. EHR extractions), we included patients who had clinically suspected COVID-19, as the currently available assays still have limited sensitivity. Technicians were employed to extract blood out of auto-analyser machines to get the earliest possible sample, enabling recruitment of patients after admission or even as outpatients. Missing data was therefore rare, and only when blood samples were simply not available, again, maximising the completeness of the cohort.

### Limitations

The major limitation of this study is the limited sample size, leading to imprecise estimates of biomarker performance. A second limitation is the composite outcome of non-invasive ventilation, ITU admission and death which has been used in major interventional COVID-19 trials. Although this is clinically useful and may aid differentiation of those who require specialist care, the provision and use of non-invasive ventilation is more clinician and hospital dependent and may be harder to extrapolate from. Finally, although around 80% of patients had blood available, 36 patients did not have stored blood, so only the routinely performed biomarkers (CRP, neutrophils, lymphocytes, neutrophil:lymphocyte ratio) are recorded for those patients.

Another potential limitation is the composite outcome. Combining non-invasive ventilation, intensive care admission and death was a deliberate choice to reflect situations requiring high-level medical care, and is clinically useful, but is more subjective than other outcomes. However, our secondary analysis of intensive care admission and death was largely similar, supporting this approach.

## Conclusion

To our knowledge this is the largest recruited UK cohort of consecutive patients presenting with COVID-19. Blood biomarkers, when performed on admission bloods, had only moderate predictive value for COVID-19, similar to age and routine clinical scores (e.g. NEWS score). IL-6 and suPAR had the best performance, and further, large prospective studies should validate the additional value of these biomarkers to routinely collected clinical information.

## Data sharing

Although we are unable to share raw data, the analytic code is available at https://github.com/gushamilton/discover_prediction/

## Data Availability

Data is not available, but code is at:
https://github.com/gushamilton/discover_prediction

## Funding

The DISCOVER study was supported by grants from the Southmead Hospital Charity and Elizabeth Blackwell Institute.

DTA is funded by a National Institute for Health Research (NIHR) Doctoral Research Fellowship (DRF-2018-11-ST2-065). The views expressed are those of the author(s) and not necessarily those of the NHS, the NIHR or the Department of Health and Social Care.

FH is funded via the National Institute for Health Academic Clinical Fellowship scheme.

The suPARnostic™ ELISA kits and IL-6/KL-6 assays were gifts from ViroGates (Birkeroed, Denmark) and Fujirebio Europe respectively for unrestricted research activity.

